# Evolving on Wheels: One-Year of Mobility Evolution in Adults with Autosomal Recessive Spastic Ataxia of Charlevoix-Saguenay

**DOI:** 10.1101/2025.09.16.25335887

**Authors:** François Routhier, Krista L. Best, Rose Gagnon, Caroline Rahn, Cynthia Gagnon, Luc J. Hébert, Isabelle Lessard, Xavier Rodrigue

**Affiliations:** School of Rehabilitation Sciences, Faculty of Medicine, Université Laval, 1050 Avenue de la Médecine, Quebec City, QC, G1V 0A6, Canada; Centre for Interdisciplinary Research in Rehabilitation and Social Integration (Cirris), Centre intégré universitaire de santé et de services sociaux (CIUSSS) de la Capitale-Nationale, 525 Wilfrid-Hamel Boulevard, Quebec City, QC, G1M 2S8, Canada; Population Health and Optimal Health Practices Axis, CHU de Québec – Université Laval Research Centre, 2705 Laurier Boulevard, Quebec City, QC, G1V 4G2, Canada; Institut de réadaptation en déficience physique de Québec (IRDPQ), Centre intégré universitaire de santé et de services sociaux (CIUSSS) de la Capitale-Nationale, 525 Wilfrid-Hamel Boulevard, Quebec City, QC, G1M 2S8, Canada; School of Rehabilitation, Faculty of Medicine and Health Sciences, Université de Sherbrooke, 3001 12th Avenue North, Sherbrooke, QC, J1H 5N4, Canada; Interdisciplinary Research Group on Neuromuscular Disorders, Hôpital de Jonquière, Centre intégré universitaire de santé et de services sociaux (CIUSSS) du Saguenay-Lac-Saint-Jean, 2230 de l’Hôpital, Jonquière, QC, G7X 7X2, Canada; Department of Radiology and Nuclear Medicine, Faculty of Medicine, Université Laval, 1050 Avenue de la Médecine, Quebec City, QC, G1V 0A6, Canada; Centre d’Étude des Conditions de vie et des Besoins de la population (ÉCOBES) – Recherche et transfert, Cégep de Jonquière, 2505 St-Hubert Street, Jonquière, QC, G7X 7W2, Canada

**Keywords:** ARSACS, manual wheelchair, actimetry, wheelchair skills, self-efficacy, mobility

## Abstract

**Introduction:** Autosomal recessive spastic ataxia of Charlevoix-Saguenay (ARSACS) is a progressive neurological disorder that leads to significant motor impairments and eventual wheelchair use in adulthood. Although previous studies have described limited mobility and participation in adult wheelchair users with ARSACS, data remains scarce. This study aimed to describe mobility evolution over one year of adult manual wheelchair users with ARSACS.

**Methods:** Longitudinal descriptive study including manual wheelchair users with genetically confirmed ARSACS (aged 18-59) recruited from two neuromuscular clinics in the province of Quebec (Canada). Participants completed validated questionnaires assessing wheelchair skills and self-efficacy, and wore two accelerometers for seven days to measure objective mobility (self-propulsion and non-propulsion). Data were collected at baseline and one year later, and analyzed using descriptive statistics and nonparametric repeated measures ANOVA.

**Results:** Thirty-four adult manual wheelchair users with ARSACS participated in the study, with a mean age of 46.1 years (± 8.3 years) and moderate to severe disease severity at baseline. Participants reported low manual wheelchair skills, but high self-efficacy regarding acquired skills. Over one year, objective mobility measures showed a non-significant decrease in distance traveled, number of bouts and bout length. Differences in objective mobility measures according to age and sex were observed, but were non-statistically significant.

**Conclusion:** This study represents a first step towards a better understanding of wheelchair mobility of adults with ARSACS. These findings have the potential to help improve their rehabilitation process.

**Highlights:**

- Manual wheelchair users reported low wheelchair skills, but high self-efficacy
- Total distance traveled, number of bouts, and bout length decreased over one year
- Proportion of self-propelled objective mobility decreased over one year
- Differences according to age and sex were observed, but were non-significant

## 1.1. Introduction

Autosomal recessive spastic ataxia of Charlevoix-Saguenay (ARSACS) is a slowly progressive neurological disorder characterized by cerebellar, pyramidal and neuropathic impairments [1, 2]. ARSACS was originally described and is still most prevalent in the Charlevoix and Saguenay-Lac-Saint-Jean regions in Quebec, Canada [1, 3]. However, cases have now been reported in many other countries, leading to its recognition as one of the most common forms of recessive spastic ataxia worldwide [1, 4, 5]. Despite individual variability in mutations and manifestations [2, 6-8], common signs include lack of coordination, distal weakness, and impaired dexterity and balance [8-11]. Gait abnormalities or falls are often the initial reasons for consultation during childhood (3.4 years on average) [9]. The evolution of these limitations will largely determine the timing of wheelchair acquisition, which occurs at a mean age of 39 years (range of 17-58 years) [8, 12]. In the province of Quebec, Canada, around 45% of adults with ARSACS are permanent wheelchair users [2, 8, 13].

Nevertheless, literature on adult wheelchair users with ARSACS or their mobility is scarce. Preliminary evidence indicates that loss of the ability to walk is associated with decreased social participation, functional independence, and motor performance in this population [8, 10]. Only one recent descriptive study by Bourassa et al. has specifically described the characteristics of adult wheelchair users with ARSACS [14]. Limited wheelchair skills, significantly impaired motor function, and restricted social participation that generally decreased with age were found. This study also provided the first data on wheelchair mobility and wheelchair life-space of adults with ARSACS. Participants reported relatively limited travel in mobility areas with their wheelchair compared to other populations, such as older manual wheelchair users [15]. However, further investigation is needed, as no objective measure of wheelchair displacement was taken.

Therefore, objectively measuring manual wheelchair mobility in adults with ARSACS is of interest. Such empirical evidence may help us better understand the profile and clinical evolution of this understudied subpopulation, an essential step to advance trial readiness [16] and to guide their rehabilitation process. It may also confirm and offer possible explanations related to the limitations in social participation and wheelchair mobility that have been documented subjectively in previous studies [8, 14, 17]. To achieve this, activity monitors (e.g., accelerometers, data loggers) have become an increasingly common method to objectively measure manual wheelchair displacement [18, 19].

Thus, the general objective of this study was to describe the mobility evolution over one year of manual wheelchair users with ARSACS. More specifically, we measured the evolution of their wheelchair skills, wheelchair self-efficacy, and objective mobility.

## 1.2. Material and Methods

### 1.2.1. Study Design and Setting

This study used a longitudinal descriptive design including comparative analyses. Data were collected as part of a larger multicenter longitudinal cohort study on the natural history of adults with ARSACS. Testing took place at the Saguenay Neuromuscular clinic (*Centre intégré universitaire de santé et de services sociaux du Saguenay-Lac-Saint-Jean*) in Saguenay (Canada) and the *Centre intégré universitaire de santé et de services sociaux de la Capitale-Nationale* in Quebec City (Canada). Ethical approval was obtained for each site through the *Centre intégré universitaire de santé et de services sociaux de la Capitale-Nationale* Research Ethics Board in rehabilitation and social integration and the *Centre intégré universitaire de santé et de services sociaux du Saguenay-Lac-Saint-Jean* Research Ethics Board. All participants provided written informed consent.

### 1.2.2. Participants and Sampling

Participants were recruited in Saguenay and Quebec City through the registry of patients with recessive ataxia from each clinic. Individuals who had participated in a previous phase of the longitudinal study were contacted again by the research team and additional participants were recruited. For all participants of the larger study, a random sampling strategy stratified by age group (14–17; 18–29; 30–39; 40–49; 50–59 years) and sex (men, women) was used. To be included, participants had to be between 18 and 59 years of age, be a manual wheelchair user (occasionally or regularly), and have a genetically confirmed ARSACS diagnosis. Participants were excluded if they had other medical conditions leading to significant functional limitations or multiple morbidities preventing them from completing all assessments.

### 1.2.3. Main Outcome Measures

#### 1.2.3.1. Questionnaires and Rating Scales

As part of the larger study, several measures (French versions) were collected, which allowed for the description of the clinical progression of the adults recruited, as well as the progression of their manual wheelchair skills and self-efficacy. A demographic questionnaire was also used to obtain data on participants’ sex, age, main occupation, level of education, and living environment.

##### Wheelchair skills and self-efficacy

Wheelchair skills and self-efficacy were quantified using the Wheelchair Skills Test Questionnaire (WST-Q-F) 5.0 and the Wheelchair Use Confidence Scale (WheelCon-F) Short Form. The WST-Q-F assesses capacity, confidence, performance, and training goals regarding 32 manual wheelchair skills. Higher percentage scores for each subscale indicate better wheelchair skills [20, 21]. The WheelCon-F includes 21 items, divided in two subscales (mobility self-efficacy and self-management efficacy), for which the respondents rate their level of confidence on a scale of 0-10 [22]. Both questionnaires have been validated among adults with ARSACS [17].

##### Motor performance

The severity of cerebellar ataxia was quantified using the Scale for the Assessment and Rating of Ataxia (SARA) [23]. The SARA includes eight items for a total score ranging from 0 (no ataxia) to 40 (severe ataxia). The Disease Severity Index for adults with ARSACS (DSI-ARSACS) was also used to measure disease-specific severity. The DSI-ARSACS assesses cerebellar, neuropathic and pyramidal ARSACS impairments through eight items [24]. A higher score (/38) indicates a more severe disease stage. Both assessments have been validated in adults with ARSACS [24, 25].

#### 1.2.3.2. Objective Mobility

Two ActiGraph wGT3X-BT accelerometers (ActiGraph Corp., Peniscola, FL) were used to measure objective mobility (self-propelled and non-propelled). The ActiGraph wGT3X-BT is a small (4.6cm x 3.3cm x 1.5cm) and lightweight (19 grams) device that can be worn on the arm and does not impede arm movements. It contains a solid state three-axis microelectromechanical system-based accelerometer with a sample rate of 30-110 Hertz and a dynamic range of 8G (https://www.actigraphcorp.com/). The validity and reliability of this device for manual wheelchair users has been demonstrated [26-29]. Activity counts measured using actigraphs were sampled at a frequency of 30 Hz (every 1/30^th^ of a second). The sampling unit (epochs) was then converted to 1s to facilitate data analysis and to ensure enough sensitivity for low-intensity activities.

Participants wore two ActiGraph accelerometers for seven consecutive days, except during sleep and hygiene periods. The first accelerometer was worn between the elbow and shoulder on their non-dominant arm using an armband. The non-dominant arm was selected, as wearing the ActiGraph on the dominant arm may result in an overestimation of activity due to extraneous arm movement [27]. The other accelerometer was attached close to the axis of the wheelchair rear wheel on the same side using a sturdy plastic case and tie-wraps. We made sure that the middle of the device was aligned parallel to a spoke of the wheel and that no undesired movement was allowed. Research suggests that using two devices is a particularly promising way of capturing self-propelled wheelchair driving in real-life environments [29-31]. More precisely, the combination of two ActiGraph GT3X-BT accelerometers has been shown to validly differentiate between self-propelled and non-propelled wheelchair driving (overall agreement of 85%) among manual wheelchair users with spinal cord injury [29]. Participants were asked to report in writing any omissions related to wearing the accelerometers (logbook), if applicable. Objective wheelchair mobility measurements taken using actigraphs were processed using a custom algorithm previously developed and validated by our research team, which allows to derive several measures of manual wheelchair physical activity (i.e., total distance traveled, number of bouts, bout length, bout speed, and maximum speed), and to discriminate between self-propulsion and non-propulsion. More details on the algorithm’s functioning can be found in a previous article by Gagnon et al. [32].

### 1.2.4. Procedures

Baseline outcome measures were administered over three sessions: one 1-h session by phone and two half-day sessions in clinic (**Figure 1**). The ActiGraph accelerometers were presented and installed during the first session in clinic. They were taken back during the second session, after the 7-day wearing period. All participants who completed the baseline assessment were subsequently contacted after one year to repeat the two measurement sessions (i.e., ARSACS rating scales, wheelchair skills and motor performance questionnaires, and 7-day actigraphs wearing period).

**Figure 1.**
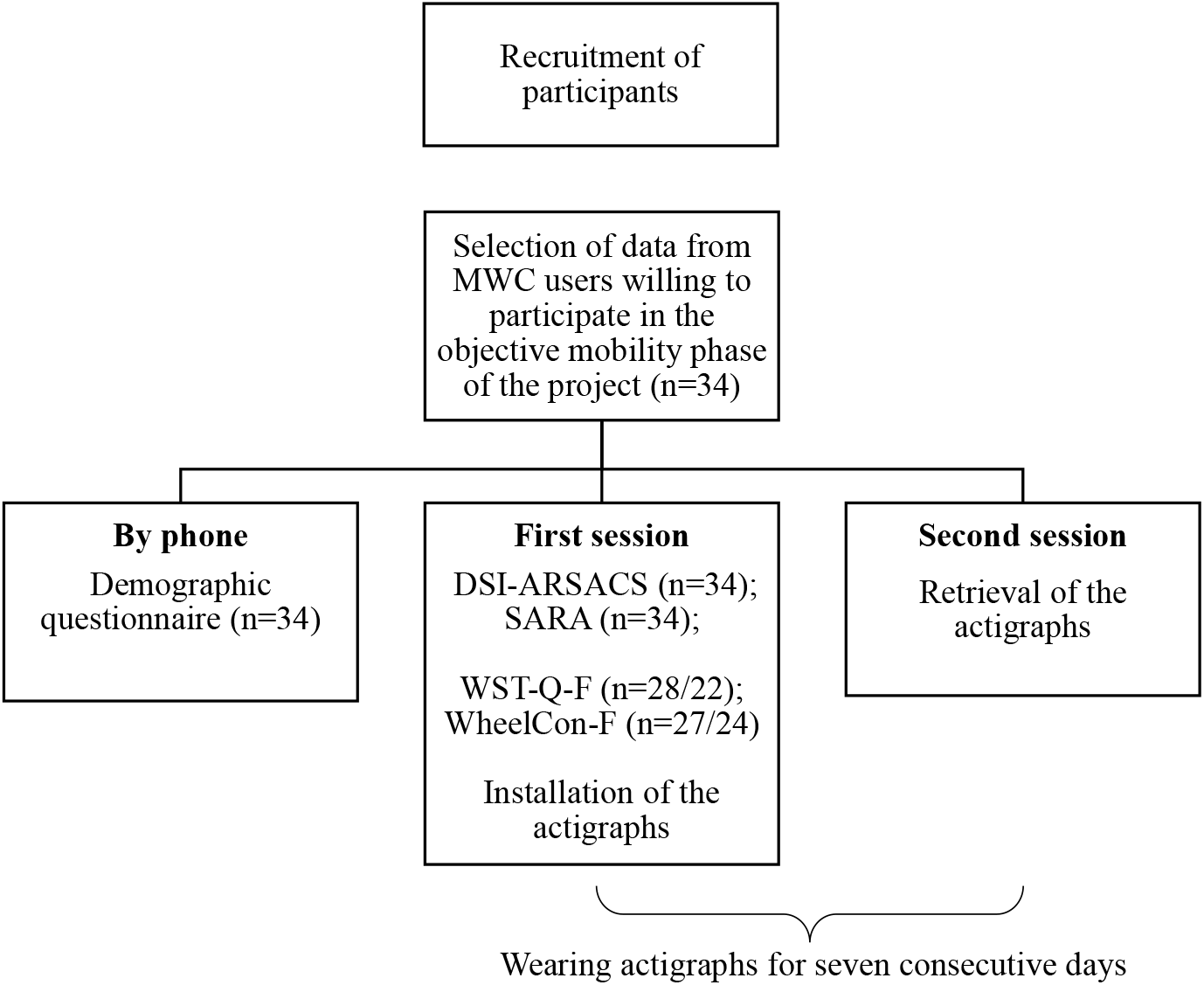
Flow diagram representing the study’s procedures and completed assessment tools during each session. MWC, manual wheelchair; DSI-ARSACS, Disease Severity Index for adults with ARSACS; SARA, Scale for the Assessment and Rating of Ataxia; WST-Q-F, Wheelchair Skills Test Questionnaire – French version; WheelCon-F, Wheelchair Use Confidence Scale – French version.

### 1.2.5. Data Analysis

Descriptive statistics (mean, standard deviation, proportion) were used to summarize participants’ sociodemographic characteristics, wheelchair skills, wheelchair self-efficacy, motor performance, and objective mobility (self-propelled and non-propelled). Objective mobility measures measured during the 7-day wearing period were separated into four measurement types to better detail possible variations in mobility habits (i.e., three days, seven days, weekend, week). In addition to analyses including all participants, stratified analyses according to age and biological sex were also performed to explore whether these two characteristics had a differential impact on the clinical evolution observed. Participants’ age was dichotomized based on the median age of the sample (≤ 46 years and ≥ 47 years). This choice was made to obtain two groups of similar size for statistical analysis purposes. Data were analyzed using nonparametric repeated measures analysis of variance (ANOVA, nparLD package). The level of statistical significance (α) was set at .05. All analyses were performed using R statistical software (versions 4.3.2 and 4.4.2, R Foundation).

## 1.3. Results

### 1.3.1. Participants’ Characteristics

**Table 1** presents the sociodemographic characteristics of the 34 adult manual wheelchair users included in the study. Mean participants’ age at baseline was 46.1 years, and 58.8% were males. Mean baseline scores for the SARA and DSI-ARSACS were respectively 27.5 ± 5.2 and 23.5 ± 4.4, indicating a significant level of ataxia (SARA) and a more severe disease stage (DSI-ARSACS). More than 70% of participants used a manual wheelchair for indoor (73.5%) and outdoor mobility (76.5%) at baseline, compared to 58.8% (indoor) and 52.9% (outdoor) at the 1-year follow-up. Participants’ autonomy level ranged from autonomous (baseline: 38.2%, 1-year follow-up: 11.8%) to complete assistance (baseline: 0.0%, 1-year follow-up: 2.9%), with the majority being partially assisted at the end of follow-up (baseline: 61.8%, 1-year follow-up: 70.6%).

**Table 1.** Participants sociodemographic characteristics (n = 34)

| Variable | Value |  |
| --- | --- | --- |
|  | Baseline | Follow-Up |
| Mean (SD) |  |  |
| Age (years) | 46.1 (8.3) | 46.8 (8.1) |
| SARA, /40 <sup>a</sup> | 27.5 (5.2) | 28.6 (4.9) |
| DSI – ARSACS, /38 <sup>b</sup> | 23.5 (4.4) | 24.8 (4.6) |
| n (%) |  |  |
| Biological sex |  |  |
| Male | 20 (58.8) |  |
| Female | 14 (41.2) |  |
| Education |  |  |
| Less than high school | 4 (11.8) |  |
| High-school degree | 10 (29.4) |  |
| Post-secondary education | 18 (52.9) |  |
| Prefer not to answer | 2 (5.9) |  |
| Deceased | 0 (0.0) | 3 (8.8) |
| Indoor mobility level <sup>c</sup> |  |  |
| Walking without help | 1 (2.9) | 0 (0.0) |
| Cane or walker | 7 (20.6) | 5 (14.7) |
| Manual wheelchair | 25 (73.5) | 20 (58.8) |
| Powered wheelchair | 0 (0.0) | 4 (11.8) |
| Both | 1 (2.9) | 0 (0.0) |
| Outdoor mobility level <sup>c</sup> |  |  |
| Walking without help | 0 (0.0) | 0 (0.0) |
| Cane or walker | 1 (2.9) | 0 (0.0) |
| Manual wheelchair | 26 (76.5) | 18 (52.9) |
| Powered wheelchair | 7 (20.6) | 8 (23.5) |
| Four-wheeled scooter | 0 (0.0) | 3 (8.8) |
| Autonomy level <sup>c</sup> |  |  |
| Autonomous | 13 (38.2) | 4 (11.8) |
| Partial assistance | 21 (61.8) | 24 (70.6) |
| Complete assistance | 0 (0.0) | 1 (2.9) |
| Main occupation <sup>c</sup> |  |  |
| Working full-time | 6 (17.6) | 4 (11.8) |
| Working part-time | 1 (2.9) | 2 (5.9) |
| Unemployed for health reasons | 22 (64.7) | 17 (50.0) |
| Other | 5 (14.7) | 6 (17.5) |
| Living arrangement <sup>c</sup> |  |  |
| Alone | 11 (32.4) | 12 (35.3) |
| Not alone | 14 (41.2) | 11 (32.4) |
| With service | 5 (14.7) | 4 (11.8) |
| Nursing home | 4 (11.8) | 2 (5.9) |

SD, Standard deviation; %, Percentage.
<sup>a</sup> Scale for the Assessment and Rating of Ataxia (SARA); total score: 0 (no ataxia) to 40 (most severe ataxia)
<sup>b</sup> Disease Severity Index for adults with Autosomal Recessive Spastic Ataxia of Charlevoix-Saguenay (ARSACS); total score 0 (no ataxia) to 38 (more severe disease stage)
<sup>c</sup> Missing data – Baseline: 0 (0.0%), Follow-Up: 5 (14.7%)

### 1.3.2. Wheelchair Skills and Self-efficacy

**Table 2** presents the mean wheelchair skills scores for the entire sample and stratified according to age and sex. Mean scores for all WST-Q-F subscales were low, ranging from 31.6 to 53.5 for the capacity subtotal, 42.3 and 64.0 for the confidence subtotal, and 31.6 and 48.1 for the performance subtotal. Scores were generally lower among participants aged 47 and older and among women, but observed differences were not statistically significant. Included participants expressed a desire to train several wheelchair abilities, both at baseline and at the 1-year follow-up (Training goals, baseline: 58.0/100, 1-year follow-up: 58.8/100). Despite low WST-Q-F scores, participants expressed a high self-efficacy level in their abilities to use a manual wheelchair (WheelCon-M-F, baseline: 7.0/10, 1-year follow-up: 7.1/10). Observed self-efficacy level was lower among users aged 47 and older, but not statistically different.

**Table 2.**
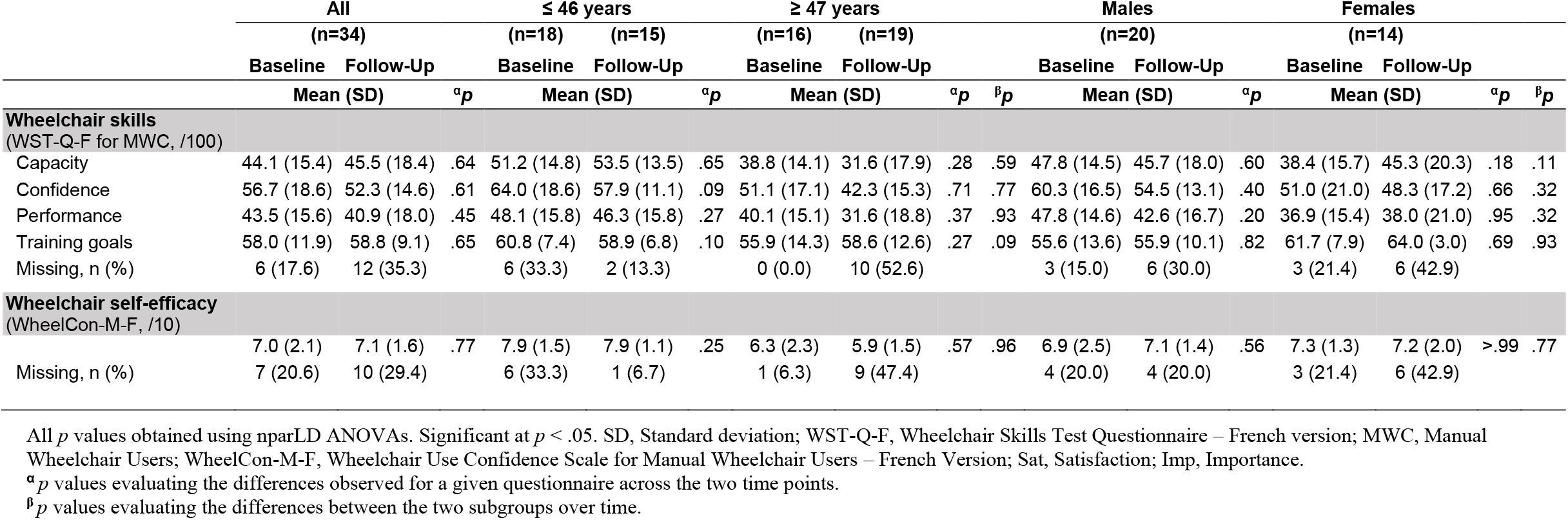
Mean wheelchair skills and self-efficacy scores and comparison between age groups and sex.

### 1.3.3. Objective mobility

**Table 3** reports the various three-day objective wheelchair mobility measures obtained using the previously validated algorithm. The seven-day, weekend, and weekly objective wheelchair mobility measures can be found in **Tables S1** to **S3** (Supplementary Material). After one year, mean total distance traveled (daily), number of bouts and bout length all decreased (non-statistically significant differences). These same measures were generally lower among participants aged 47 and older (when compared to those aged 46 and younger) and among women (when compared to men); however, these differences were not significant. Moreover, the proportion of the mean total distance traveled and number of bouts made without self-propelling increased between the initial assessment and 1-year follow-up (p>.05). Measures of bout speed and maximum speed remained generally similar, regardless of age, sex, and objective wheelchair mobility measurement type (three days, seven days, weekend, week).

**Table 3.**
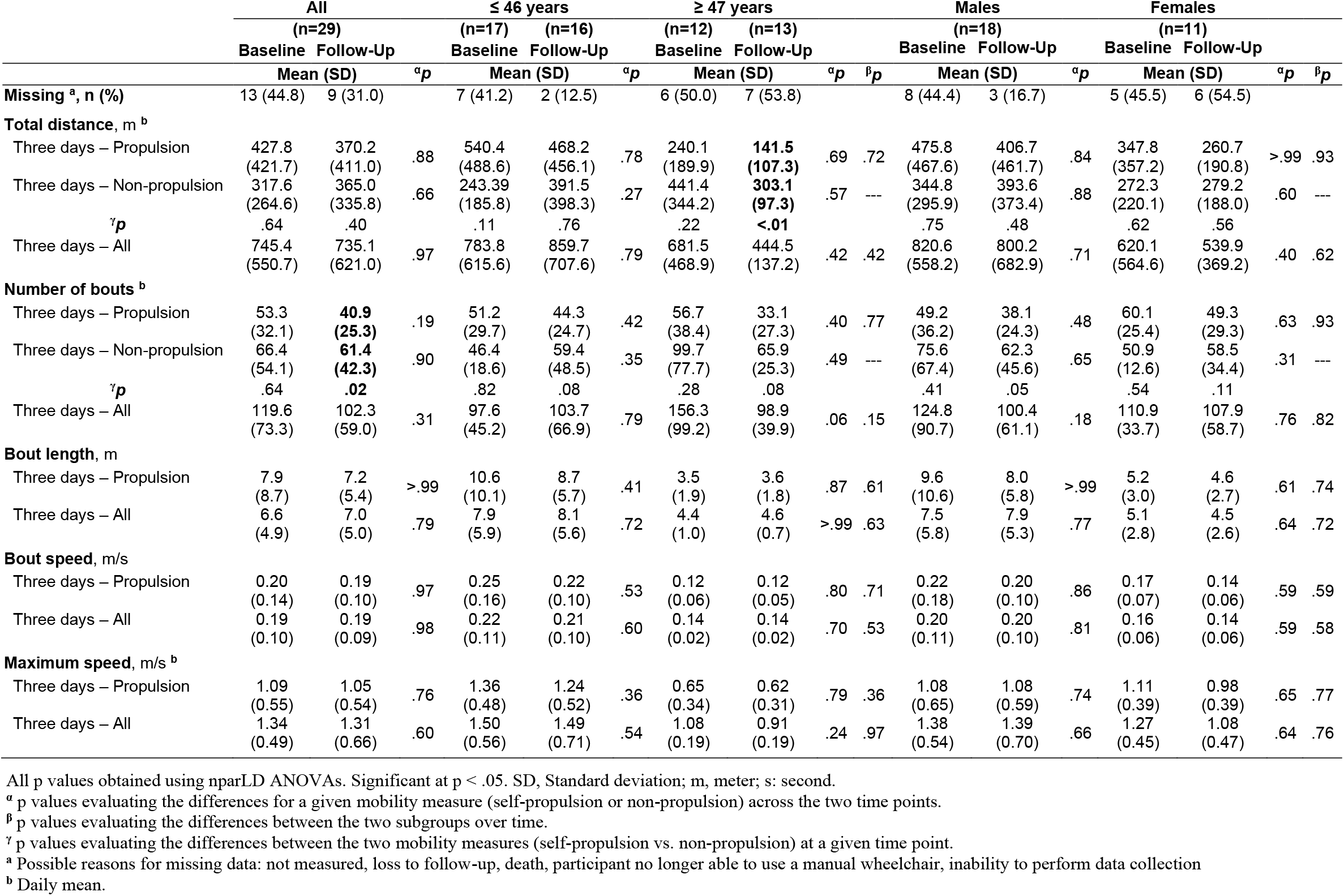
Mean wheelchair objective mobility measures - all and propulsion only - and comparison at a given time point (three days)

## 1.4. Discussion

Mean wheelchair skills scores obtained by participants via the WST-Q-F were low, both at baseline and at the 1-year follow-up. Although no normative data or recognized cut-offs for manual wheelchair skills exist, mean observed scores for the capacity, confidence, and performance subscales were lower than those reported in the scientific literature for manual wheelchair users presenting with a variety of neurological conditions [20, 33]. Several studies report a significant positive association between WST-Q scores and cardiovascular health [34, 35]. High WST-Q scores are also associated with increased participation in daily life and social activities, greater confidence, independence, and basic mobility, all of which contribute to cardiovascular health and quality of life [20, 36]. It would thus be of interest in subsequent studies to evaluate the impact of targeted interventions aimed at improving the various skills listed in the WST-Q (e.g., gets over obstacle, ascends/descends high curb) during the rehabilitation process of manual wheelchair users with ARSACS, especially considering the degenerative nature of this neurological condition [8]. Despite low wheelchair skills scores, recruited participants presented high overall WheelCon-M scores, reflecting a high self-efficacy towards their manual wheelchair skills. These high scores could be due to the fact that adults with ARSACS have low expectations regarding the use of a manual wheelchair, thereby increasing their self-efficacy with regards to the skills they have already mastered [14].

Objective wheelchair mobility measurements showed that participants presented low levels of objective mobility at baseline, and that these levels tended to decrease after one year. Previous studies have reported that adult manual wheelchair users with a neurological condition tend to travel between 1.4 and 2.5 kilometers per day [37, 38], which is much higher than the mean total daily distance measured at baseline (745.4 ± 550.7 meters) and follow-up (735.1 ± 621.0 meters). The proportion of bouts and total daily distance travelled by self-propulsion also tended to decrease over time, especially among participants aged 47 years and older (1-year follow-up propulsion vs. other: total distance, p<.01; number of bouts, p=.08). These results are important, in that manual wheelchair users who travel greater distance each day tend to be more physically active and engage in more intense physical activities [39, 40]. Greater total daily distance is also associated with improved function [41], lower depression scores, and greater life satisfaction [42]. Special attention should thus be given to interventions that promote mobility in adult manual wheelchair users with ARSACS, given the higher risk of manual wheelchair users developing comorbidities when exhibiting higher levels of physical inactivity [43].

Recruited participants expressed a significant need for training, with scores of 58.0% (baseline) and 58.8% (1-year follow-up) on the WST-Q-F’s training goals subscale. This finding is consistent with other studies indicating that manual wheelchair users may not receive enough training to improve their wheelchair skills [44, 45]. When given the opportunity to receive training, such training is often reported to be inconsistent, not tailored to individual patients’ needs, and not addressing advanced skills and environmental adaptation [44, 45]. The training provided also tend to focus on the acquisition of short-term skills, neglecting long-term confidence, self-efficacy, and community participation [44, 46]. Some preliminary studies report that engaging in physical activity may help slow the progression of certain neurological diseases [47], including ARSACS [48]. Greater attention should be given in the coming years to implementing and validating manual wheelchair skills programs that are consistent, tailored to the individual needs of adults with ARSACS, and which include advanced skills training. Such programs could increase the mobility level of adults living with this condition, and foster greater community participation.

### 1.4.1. Strengths and Limitations

Several limitations must be considered when interpreting the results of this study. The analyses performed should be regarded as exploratory given the small number of participants included. Furthermore, various reasons, such as loss to follow-up, death, loss of ability to use a manual wheelchair, and incapacity to perform the actigraphy protocol, led to a relatively high rate of missing data, both during the initial data collection and 1-year follow-up. In future studies, it will be important to ensure that the actigraphy protocol can be easily performed by participants, especially considering the high fatigability of persons with ARSACS and, more broadly, of any adult manual wheelchair user with a neurological condition. It is also possible that the placement of the actigraphs on the wheel of the manual wheelchair and on the non-dominant arm did not capture movements performed using different self-propulsion patterns, such as dominant arm and one foot or two feet. The objective mobility measurements reported in this study should therefore be viewed as minimums.

Despite these limitations, we believe that this study has some noteworthy strengths. It comprehensively reports numerous preliminary longitudinal objective mobility measures that may help better understand the profile and clinical evolution of this understudied population, representing an essential step toward future clinical trials and tailoring of their rehabilitation services. Actigraphy data reported in this study were derived using a previously validated conversion algorithm with excellent accuracy. Finally, use of this same algorithm allowed to explore the proportion of self-propulsion for certain objective mobility measures (i.e., total distance, number of bouts). These mobility measures pave the way for future studies, such as correlation studies focusing on the most significant factors contributing to the observed decline in wheelchair mobility.

## 1.5. Conclusion

Results of this study indicate that adults with ARSACS present limited manual wheelchair skills, high wheelchair self-efficacy, and low levels of objective mobility. In addition, total objective mobility done while self-propelling tended to decrease after one year. This study represents a first step towards a better understanding of mobility levels of adults with ARSACS. These findings have the potential to help improve their rehabilitation process.

## Data Availability

All data produced in the present study are available upon reasonable request to the authors.

## 1.6. Acknowledgements

The Authors would like to thank the following persons for their contributions: all members of the research team who contributed to data collection, and Julie Bourassa, Alexandre Desgagné-Lebeuf, Valérie Moisan, Jean Leblond, and Bernard Brais for their contribution to the study’s conception, data collection and analysis.

## 1.7. Statements and declarations

### 1.7.3. Authors’ Contributions

Conceptualization: FR, KLB, CG, LJH, IL, XR; Funding acquisition: FR, KLB, CG, LJH, XR; Investigation: FR, KLB, CR, CG; Formal analysis: FR, KLB, RG; Visualization: FR, KLB, RG; Writing – original draft: FR, KLB, RG; Writing – review and editing: FR, KLB, RG, CR, CG, LJH, IL, XR.

## References

[1] S. Vermeer, B. van de Warrenburg, E. Kamsteeg, B. Brais, M.A. Synofzik, Dec 9 [Updated 2020 Jan 2], GeneReviews®[Internet]. Seattle (WA): University of Washington, Seattle 2024 (1993).

[2] M.M. Briand, X. Rodrigue, I. Lessard, J. Mathieu, B. Brais, I. Cote, C. Gagnon, Expanding the clinical description of autosomal recessive spastic ataxia of Charlevoix-Saguenay, J Neurol Sci 400 (2019) 39–41.

[3] J.P. Bouchard, A. Barbeau, R. Bouchard, R.W. Bouchard, Autosomal Recessive Spastic Ataxia of Charlevoix-Saguenay, Canadian Journal of Neurological Sciences / Journal Canadien des Sciences Neurologiques 5(1) (2015) 61–69.

[4] C.M. Gomez, ARSACS goes global, Lippincott Williams & Wilkins, 2004, pp. 10–11.

[5] Y. Bouhlal, R. Amouri, G. El Euch-Fayeche, F. Hentati, Autosomal recessive spastic ataxia of Charlevoix-Saguenay: an overview, Parkinsonism Relat Disord 17(6) (2011) 418–22.

[6] R. Lariviere, R. Gaudet, B.J. Gentil, M. Girard, T.C. Conte, S. Minotti, K. Leclerc-Desaulniers, K. Gehring, R.A. McKinney, E.A. Shoubridge, P.S. McPherson, H.D. Durham, B. Brais, Sacs knockout mice present pathophysiological defects underlying autosomal recessive spastic ataxia of Charlevoix-Saguenay, Hum Mol Genet 24(3) (2015) 727–39.

[7] I. Thiffault, M.J. Dicaire, M. Tetreault, K.N. Huang, J. Demers-Lamarche, G. Bernard, A. Duquette, R. Lariviere, K. Gehring, A. Montpetit, P.S. McPherson, A. Richter, L. Montermini, J. Mercier, G.A. Mitchell, N. Dupre, C. Prevost, J.P. Bouchard, J. Mathieu, B. Brais, Diversity of ARSACS mutations in French-Canadians, Can J Neurol Sci 40(1) (2013) 61–6.

[8] C. Gagnon, B. Brais, I. Lessard, C. Lavoie, I. Cote, J. Mathieu, From motor performance to participation: a quantitative descriptive study in adults with autosomal recessive spastic ataxia of Charlevoix-Saguenay, Orphanet J Rare Dis 13(1) (2018) 165.

[9] A. Duquette, B. Brais, J.P. Bouchard, J. Mathieu, Clinical presentation and early evolution of spastic ataxia of Charlevoix-Saguenay, Mov Disord 28(14) (2013) 2011–4.

[10] C. Gagnon, J. Desrosiers, J. Mathieu, Autosomal recessive spastic ataxia of Charlevoix-Saguenay: upper extremity aptitudes, functional independence and social participation, International Journal of Rehabilitation Research 27(3) (2004) 253–256.

[11] C. Gagnon, I. Lessard, C. Lavoie, I. Cote, R. St-Gelais, J. Mathieu, B. Brais, An exploratory natural history of ataxia of Charlevoix-Saguenay: A 2-year follow-up, Neurology 91(14) (2018) e1307–e1311.

[12] J.-P. Bouchard, Recessive spastic ataxia of Charlevoix-Saguenay, Handbook of clinical neurology; Hereditary neuropathy and spinocerebellar atrophies 60 (1991) 451–459.

[13] I. Lessard, B. Brais, I. Cote, C. Lavoie, M. Synofzik, J. Mathieu, C. Gagnon, Assessing mobility and balance in Autosomal Recessive Spastic Ataxia of Charlevoix-Saguenay population: Validity and reliability of four outcome measures, J Neurol Sci 390 (2018) 4–9.

[14] J. Bourassa, F. Routhier, C. Gagnon, C. Rahn, L.J. Hebert, R. St-Gelais, X. Rodrigue, B. Brais, K.L. Best, Wheelchair mobility, motor performance and participation of adult wheelchair users with ARSACS: a cross-sectional study, Disabil Rehabil Assist Technol 18(4) (2023) 378–386.

[15] B.M. Sakakibara, W.C. Miller, J.J. Eng, C.L. Backman, F. Routhier, Influences of wheelchair-related efficacy on life-space mobility in adults who use a wheelchair and live in the community, Phys Ther 94(11) (2014) 1604–13.

[16] Center for Drug Evaluation and Research, Roadmap to patient-focused outcome measurement in clinical trials, 2019. https://www.fda.gov/drugs/drug-development-tool-ddt-qualification-programs/roadmap-patient-focused-outcome-measurement-clinical-trials-text-version. (Accessed 2025 August 30.

[17] J. Bourassa, K.L. Best, C. Gagnon, L.J. Hebert, B. Brais, F. Routhier, Measurement properties of wheelchair use assessment tools in adults with autosomal recessive spastic ataxia of Charlevoix-Saguenay, Disabil Rehabil Assist Technol 17(8) (2022) 907–915.

[18] F. Routhier, J. Lettre, W.C. Miller, J.F. Borisoff, K. Keetch, I.M. Mitchell, C. Research Team, Data logger technologies for manual wheelchairs: A scoping review, Assist Technol 30(2) (2018) 51–58.

[19] K. Tsang, B.E. Dicianno, Validity of activity monitors in wheelchair users: A systematic review, Journal of Rehabilitation Research and Development 53(6) (2016) 641.

[20] R.L. Kirby, L.A. Worobey, R. Cowan, J.P. Pedersen, A.W. Heinemann, T.A. Dyson-Hudson, M. Shea, C. Smith, P.W. Rushton, M.L. Boninger, Wheelchair Skills Capacity and Performance of Manual Wheelchair Users With Spinal Cord Injury, Arch Phys Med Rehabil 97(10) (2016) 1761–9.

[21] P.W. Rushton, R.L. Kirby, W.C. Miller, Manual wheelchair skills: objective testing versus subjective questionnaire, Arch Phys Med Rehabil 93(12) (2012) 2313–8.

[22] B.M. Sakakibara, W.C. Miller, P.W. Rushton, Rasch analyses of the wheelchair use confidence scale, Arch Phys Med Rehabil 96(6) (2015) 1036–44.

[23] T. Schmitz-Hubsch, S.T. Du Montcel, L. Baliko, J. Berciano, S. Boesch, C. Depondt, P. Giunti, C. Globas, J. Infante, J.-S. Kang, Scale for the assessment and rating of ataxia: development of a new clinical scale, Neurology 66(11) (2006) 1717–1720.

[24] C. Gagnon, B. Brais, I. Lessard, C. Lavoie, I. Cote, J. Mathieu, Development and validation of a disease severity index for ataxia of Charlevoix-Saguenay, Neurology 93(16) (2019) e1543–e1549.

[25] D. Bourcier, M. Belanger, I. Cote, B. Brais, M. Synofzik, J.D. Brisson, X. Rodrigue, M.M. Gagnon, J. Mathieu, C. Gagnon, Documenting the psychometric properties of the scale for the assessment and rating of ataxia to advance trial readiness of Autosomal Recessive Spastic Ataxia of Charlevoix-Saguenay, J Neurol Sci 417 (2020) 117050.

[26] S. Bourassa, K.L. Best, M. Racine, J. Borisoff, J. Leblond, F. Routhier, Use of actigraphy to measure real-world physical activities in manual wheelchair users, J Rehabil Assist Technol Eng 7 (2020) 2055668320907814.

[27] X. García-Massó, P. Serra-Añó, L.M. García-Raffi, E.A. Sánchez-Pérez, J. López-Pascual, L.M. Gonzalez, Validation of the use of Actigraph GT3X accelerometers to estimate energy expenditure in full time manual wheelchair users with spinal cord injury, Spinal Cord 51(12) (2013) 898–903.

[28] Y.C. Learmonth, D. Kinnett-Hopkins, I.M. Rice, J.L. Dysterheft, R.W. Motl, Accelerometer output and its association with energy expenditure during manual wheelchair propulsion, Spinal Cord 54(2) (2016) 110–4.

[29] H. Kooijmans, H.L. Horemans, H.J. Stam, J.B. Bussmann, Valid detection of self-propelled wheelchair driving with two accelerometers, Physiol Meas 35(11) (2014) 2297–306.

[30] T.E. Nightingale, P.C. Rouse, D. Thompson, J.L.J. Bilzon, Measurement of Physical Activity and Energy Expenditure in Wheelchair Users: Methods, Considerations and Future Directions, Sports Med Open 3(1) (2017) 10.

[31] D. Ding, S. Hiremath, Y. Chung, R. Cooper, Detection of wheelchair user activities using wearable sensors, International Conference on Universal Access in Human-Computer Interaction, Springer, 2011, pp. 145–152.

[32] R. Gagnon, K.L. Best, F. Routhier, Validation of an innovative two-part algorithm for detecting self-propulsion in manual wheelchair users, medRxiv (2024) 2024.11.14.24313548.

[33] P. Inkpen, K. Parker, R.L. Kirby, Manual wheelchair skills capacity versus performance, Arch Phys Med Rehabil 93(6) (2012) 1009–13.

[34] R.L. Kirby, S. de Groot, R.E. Cowan, Relationship between wheelchair skills scores and peak aerobic exercise capacity of manual wheelchair users with spinal cord injury: a cross-sectional study, Disabil Rehabil 42(1) (2020) 114–121.

[35] S.L. Silveira, S. de Groot, R.E. Cowan, Association between individual wheelchair skills and fitness in community-dwelling manual wheelchair users with spinal cord injuries, Disabil Rehabil Assist Technol 19(1) (2024) 60–65.

[36] O.J. Kilkens, M.W. Post, A.J. Dallmeijer, F.W. van Asbeck, L.H. van der Woude, Relationship between manual wheelchair skill performance and participation of persons with spinal cord injuries 1 year after discharge from inpatient rehabilitation, J Rehabil Res Dev 42(3 Suppl 1) (2005) 65–73.

[37] S.E. Sonenblum, S. Sprigle, Wheelchair use in ultra-lightweight wheelchair users, Disabil Rehabil Assist Technol 12(4) (2017) 396–401.

[38] V. Lemay, F. Routhier, L. Noreau, S.H. Phang, K.A. Ginis, Relationships between wheelchair skills, wheelchair mobility and level of injury in individuals with spinal cord injury, Spinal Cord 50(1) (2012) 37–41.

[39] M.L. Tolerico, D. Ding, R.A. Cooper, D.M. Spaeth, S.G. Fitzgerald, R. Cooper, A. Kelleher, M.L. Boninger, Assessing mobility characteristics and activity levels of manual wheelchair users, The Journal of Rehabilitation Research and Development 44(4) (2007) 561.

[40] S.E. Sonenblum, S. Sprigle, R.A. Lopez, Manual wheelchair use: bouts of mobility in everyday life, Rehabil Res Pract 2012 (2012) 753165.

[41] Y. Kimura, S. Ohji, N. Nishio, Y. Abe, H. Ogawa, R. Taguchi, Y. Otobe, M. Yamada, The impact of wheelchair propulsion based physical activity on functional recovery in stroke rehabilitation: a multicenter observational study, Disabil Rehabil 44(10) (2022) 2027–2032.

[42] S.J. Mulroy, P.E. Hatchett, V.J. Eberly, L.L. Haubert, S. Conners, J. Gronley, E. Garshick, P.S. Requejo, Objective and Self-Reported Physical Activity Measures and Their Association With Depression and Satisfaction With Life in Persons With Spinal Cord Injury, Arch Phys Med Rehabil 97(10) (2016) 1714–20.

[43] T.B. Willingham, J. Stowell, G. Collier, D. Backus, Leveraging Emerging Technologies to Expand Accessibility and Improve Precision in Rehabilitation and Exercise for People with Disabilities, International Journal of Environmental Research and Public Health 21(1) (2024) 79.

[44] K.L. Best, F. Routhier, W.C. Miller, A description of manual wheelchair skills training: current practices in Canadian rehabilitation centers, Disabil Rehabil Assist Technol 10(5) (2015) 393–400.

[45] C.T. Rusek, M. Kleven, C. Walker, K. Walker, R. Heeb, K.A. Morgan, Perspectives of inpatient rehabilitation clinicians on the state of manual wheelchair training: a qualitative analysis, Disabil Rehabil Assist Technol 18(7) (2023) 1154–1162.

[46] K. Charlton, C. Murray, N. Layton, E. Ong, L. Farrar, T. Serocki, S. Attrill, Manual wheelchair training approaches and intended training outcomes for adults who are new to wheelchair use: A scoping review, Aust Occup Ther J 72(1) (2025) e12992.

[47] S. Proschinger, P. Kuhwand, A. Rademacher, D. Walzik, C. Warnke, P. Zimmer, N. Joisten, Fitness, physical activity, and exercise in multiple sclerosis: a systematic review on current evidence for interactions with disease activity and progression, J Neurol 269(6) (2022) 2922–2940.

[48] O. Audet, H.T. Bui, M. Allisse, A.S. Comtois, M. Leone, Assessment of the impact of an exercise program on the physical and functional capacity in patients with autosomal recessive spastic ataxia of Charlevoix-Saguenay: An exploratory study, Intractable Rare Dis Res 7(3) (2018) 164–171.

