## Supplementary Material for "Evolving on Wheels: One-Year of Mobility Evolution in Adults with Autosomal Recessive Spastic Ataxia of Charlevoix-Saguenay"

**Table S1. Mean wheelchair objective mobility measures – all and propulsion only – and comparison at a given time point (seven days)**

| | All | | | ≤ 46 years | | | ≥ 47 years | | | | Males | | | Females | | | |
|---|---|---|---|---|---|---|---|---|---|---|---|---|---|---|---|---|---|
| | (n=29) | | | (n=17) | (n=16) | | (n=12) | (n=13) | | | (n=18) | | | (n=11) | | | |
| | Baseline | Follow-Up | | Baseline | Follow-Up | | Baseline | Follow-Up | | | Baseline | Follow-Up | | Baseline | Follow-Up | | |
| | Mean (SD) | | $^{\alpha}p$ | Mean (SD) | | $^{\alpha}p$ | Mean (SD) | | $^{\alpha}p$ | $^{\beta}p$ | Mean (SD) | | $^{\alpha}p$ | Mean (SD) | | $^{\alpha}p$ | $^{\beta}p$ |
| **Missing [a], n (%)** | 13 (44.8) | 11 (37.9) | | 7 (41.2) | 3 (18.8) | | 6 (50.0) | 8 (61.5) | | | 8 (44.4) | 5 (27.8) | | 5 (45.5) | 6 (54.5) | | |
| **Total distance**, m [b] | | | | | | | | | | | | | | | | | |
| Seven days – Propulsion | 382.8 (352.6) | 340.2 (293.3) | .90 | 478.9 (402.2) | 417.6 (306.5) | .55 | 222.7 (180.5) | 139.0 (117.0) | >.99 | .90 | 411.1 (389.3) | 372.5 (338.7) | .94 | 335.8 (309.9) | 256.5 (96.4) | .69 | .95 |
| Seven days – Non-propulsion | 329.9 (247.7) | 321.5 (242.4) | .78 | 259.6 (150.6) | 323.4 (273.9) | .53 | 446.9 (341.8) | 316.7 (157.0) | .55 | --- | 358.3 (293.3) | 332.1 (279.6) | .59 | 282.5 (157.6) | 294.0 (118.2) | .55 | --- |
| $^{\gamma}p$ | .90 | .87 | | .37 | .60 | | .18 | .43 | | | .94 | .82 | | .69 | .11 | | |
| Seven days – All | 712.7 (476.2) | 661.7 (450.9) | .77 | 738.5 (507.7) | 741.0 (506.4) | .94 | 669.6 (461.2) | 455.7 (146.6) | .39 | .49 | 769.4 (502.7) | 704.5 (515.9) | .60 | 618.2 (456.4) | 550.5 (210.7) | .72 | .55 |
| **Number of bouts** [b] | | | | | | | | | | | | | | | | | |
| Seven days – Propulsion | 49.0 (29.8) | **37.4 (20.5)** | .23 | 48.1 (28.9) | 41.8 (18.5) | .50 | 50.5 (34.0) | 25.9 (23.0) | .51 | .59 | 44.7 (32.4) | 34.9 (20.3) | .58 | 56.1 (25.8) | **43.9 (21.6)** | .38 | .80 |
| Seven days – Non-propulsion | 66.7 (46.5) | **59.9 (41.1)** | .69 | 52.3 (21.8) | 60.4 (46.3) | .41 | 90.8 (67.2) | 58.6 (27.4) | .24 | --- | 71.8 (58.4) | 59.8 (45.6) | .43 | 58.3 (14.5) | **60.1 (30.5)** | .45 | --- |
| $^{\gamma}p$ | .21 | **.03** | | .37 | .09 | | .15 | .22 | | | .28 | .19 | | .46 | **.03** | | |
| Seven days – All | 115.7 (66.7) | 97.3 (54.2) | .32 | 100.4 (47.4) | 102.2 (60.7) | .80 | **141.2 (89.7)** | **84.5 (34.2)** | **.04** | .16 | 116.5 (81.5) | 94.7 (57.9) | .27 | 114.4 (37.3) | 104.1 (48.6) | .87 | .96 |
| **Bout length**, m | | | | | | | | | | | | | | | | | |
| Seven days – Propulsion | 7.8 (7.8) | 7.3 (5.2) | .78 | 10.3 (8.9) | 8.5 (5.6) | .39 | 3.6 (1.8) | 4.2 (2.5) | .42 | .40 | 9.3 (9.5) | 8.2 (6.0) | .87 | 5.2 (2.5) | 5.0 (1.0) | .78 | .76 |
| Seven days – All | 6.7 (4.3) | 7.0 (4.1) | .58 | 7.8 (5.1) | 7.1 (4.3) | .43 | 4.9 (1.5) | 6.8 (3.8) | .42 | .29 | 7.7 (5.1) | 7.8 (4.5) | .85 | 5.0 (2.1) | 4.9 (1.0) | .64 | .98 |
| **Bout speed**, m/s | | | | | | | | | | | | | | | | | |
| Seven days – Propulsion | 0.20 (0.14) | 0.19 (0.09) | .91 | 0.25 (0.15) | 0.21 (0.10) | .29 | 0.12 (0.05) | 0.13 (0.06) | .54 | .40 | 0.22 (0.17) | 0.20 (0.10) | .97 | 0.17 (0.06) | 0.15 (0.03) | .93 | .78 |
| Seven days – All | 0.19 (0.09) | 0.19 (0.08) | .74 | 0.21 (0.10) | 0.19 (0.08) | .30 | 0.15 (0.03) | 0.19 (0.09) | .45 | .22 | 0.21 (0.10) | 0.21 (0.09) | .81 | 0.16 (0.05) | 0.15 (0.03) | .92 | .76 |
| **Maximum speed**, m/s [b] | | | | | | | | | | | | | | | | | |
| Seven days – Propulsion | 1.15 (0.47) | 1.09 (0.54) | .48 | 1.37 (0.39) | 1.25 (0.53) | .15 | 0.78 (0.35) | 0.68 (0.31) | .69 | .56 | 1.12 (0.55) | 1.10 (0.60) | .64 | 1.20 (0.32) | 1.07 (0.38) | .46 | .75 |
| Seven days – All | 1.49 (0.46) | 1.46 (0.53) | .59 | 1.64 (0.51) | 1.52 (0.59) | .25 | 1.26 (0.27) | 1.30 (0.33) | .72 | .49 | 1.49 (0.53) | 1.54 (0.61) | .90 | 1.49 (0.38) | 1.27 (0.21) | .26 | .38 |

All p values obtained using nparLD ANOVAs. Significant at $p < .05$. SD, Standard deviation; m, meter; s: second.

$^{\alpha}$ p values evaluating the differences for a given mobility measure (self-propulsion or non-propulsion) across the two time points.

$^{\beta}$ p values evaluating the differences between the two subgroups over time.

$^{\gamma}$ p values evaluating the differences between the two mobility measures (self-propulsion vs. non-propulsion) at a given time point.

[a] Possible reasons for missing data: not measured, loss to follow-up, death, participant no longer able to use a manual wheelchair, inability to perform data collection

[b] Daily mean.

**Table S2. Mean wheelchair objective mobility measures – all and propulsion only – and comparison at a given time point (weekend)**

| | All | | | ≤ 46 years | | | ≥ 47 years | | | | Males | | | Females | | | |
|---|---|---|---|---|---|---|---|---|---|---|---|---|---|---|---|---|---|
| | (n=29) | | | (n=17) | (n=16) | | (n=12) | (n=13) | | | (n=18) | | | (n=11) | | | |
| | Baseline | Follow-Up | | Baseline | Follow-Up | | Baseline | Follow-Up | | | Baseline | Follow-Up | | Baseline | Follow-Up | | |
| | Mean (SD) | | $^{\alpha}p$ | Mean (SD) | | $^{\alpha}p$ | Mean (SD) | | $^{\alpha}p$ | $^{\beta}p$ | Mean (SD) | | $^{\alpha}p$ | Mean (SD) | | $^{\alpha}p$ | $^{\beta}p$ |
| **Missing [a], n (%)** | 13 (44.8) | 10 (34.5) | | 7 (41.2) | 3 (18.8) | | 6 (50.0) | 7 (53.8) | | | 8 (44.4) | 4 (22.2) | | 5 (45.5) | 6 (54.5) | | |
| **Total distance**, m [b] | | | | | | | | | | | | | | | | | |
| Weekend – Propulsion | 370.4 (368.6) | 351.1 (435.9) | .57 | 456.3 (428.2) | 471.3 (482.9) | .76 | 227.3 (195.1) | **90.6 (73.1)** | .12 | .36 | 392.0 (367.8) | 408.2 (497.6) | .68 | 334.5 (401.9) | **191.2 (84.2)** | >.99 | .96 |
| Weekend – Non-propulsion | 289.8 (239.9) | 311.6 (205.1) | .55 | 224.8 (185.8) | 296.6 (211.3) | .14 | 398.2 (296.6) | **344.0 (206.1)** | .64 | --- | 330.2 (285.3) | 318.3 (238.1) | .87 | **222.5 (132.0)** | **292.8 (66.4)** | **.01** | --- |
| $^{\gamma}p$ | .96 | .41 | | .29 | .76 | | .38 | **.048** | | | .93 | .96 | | >.99 | **>.01** | | |
| Weekend – All | 660.2 (514.2) | 662.7 (549.0) | .97 | 681.1 (573.7) | 768.0 (628.2) | .66 | 625.5 (445.6) | 434.5 (216.0) | .48 | .34 | 722.2 (525.9) | 726.5 (629.0) | .70 | 557.0 (524.4) | 483.9 (127.8) | .19 | .42 |
| **Number of bouts** [b] | | | | | | | | | | | | | | | | | |
| Weekend – Propulsion | 46.7 (29.8) | **38.1 (20.8)** | .62 | 44.0 (28.2) | 44.5 (18.3) | .53 | 51.1 (34.7) | **24.2 (20.4)** | .07 | .16 | 42.5 (33.2) | 37.4 (24.0) | .97 | 53.6 (24.3) | **39.9 (8.8)** | .32 | .66 |
| Weekend – Non-propulsion | 62.9 (50.2) | **64.1 (42.6)** | .72 | 46.2 (26.0) | 59.6 (45.2) | .25 | 90.8 (69.8) | **74.0 (38.2)** | .71 | --- | 69.0 (63.0) | 64.6 (46.6) | .87 | 52.8 (14.8) | **62.8 (33.4)** | .45 | --- |
| $^{\gamma}p$ | .12 | **.02** | | .32 | .13 | | .15 | **.048** | | | .08 | .10 | | >.99 | **.02** | | |
| Weekend – All | 109.6 (71.5) | 102.2 (55.8) | .90 | 90.2 (49.5) | 104.0 (61.5) | .59 | 141.8 (94.4) | 98.2 (45.8) | .38 | .31 | 111.5 (89.7) | 102.0 (62.5) | >.99 | 106.3 (28.6) | 102.7 (36.0) | .86 | .68 |
| **Bout length**, m | | | | | | | | | | | | | | | | | |
| Weekend – Propulsion | 8.9 (11.7) | 7.0 (6.1) | .73 | 12.1 (14.1) | 8.8 (6.6) | .47 | 3.6 (1.7) | 3.0 (1.6) | .59 | .86 | 11.2 (14.4) | 7.8 (6.9) | .38 | 5.2 (4.1) | 4.6 (2.1) | >.99 | .60 |
| Weekend – All | 6.9 (6.0) | 6.4 (4.0) | .52 | 8.3 (7.2) | 7.2 (4.6) | .92 | 4.4 (1.0) | 4.5 (0.7) | .78 | .63 | 8.1 (7.1) | 6.8 (4.4) | --- | 4.8 (3.0) | 5.0 (2.5) | .67 | .65 |
| **Bout speed**, m/s | | | | | | | | | | | | | | | | | |
| Weekend – Propulsion | 0.21 (0.17) | 0.18 (0.11) | .74 | 0.26 (0.19) | 0.22 (0.11) | .37 | 0.12 (0.05) | 0.11 (0.05) | .60 | .91 | 0.24 (0.20) | 0.20 (0.12) | .42 | 0.16 (0.08) | 0.15 (0.05) | .82 | .55 |
| Weekend – All | 0.19 (0.11) | 0.18 (0.07) | .64 | 0.22 (0.14) | 0.20 (0.08) | .82 | 0.14 (0.02) | 0.14 (0.01) | .63 | .62 | 0.21 (0.13) | 0.19 (0.08) | .95 | 0.15 (0.06) | 0.15 (0.05) | .68 | .68 |
| **Maximum speed**, m/s [b] | | | | | | | | | | | | | | | | | |
| Weekend – Propulsion | 0.98 (0.54) | 0.98 (0.50) | .90 | 1.18 (0.57) | 1.13 (0.44) | .61 | 0.65 (0.28) | 0.64 (0.48) | .56 | .83 | 1.02 (0.67) | 1.01 (0.57) | .69 | 0.92 (0.26) | 0.87 (0.20) | .94 | .88 |
| Weekend – All | 1.25 (0.53) | 1.31 (0.66) | .91 | 1.31 (0.63) | 1.44 (0.71) | .55 | 1.16 (0.36) | 1.04 (0.50) | .60 | .44 | 1.37 (0.58) | 1.32 (0.61) | .79 | 1.06 (0.41) | 1.30 (0.88) | .87 | .64 |

All p values obtained using nparLD ANOVAs. Significant at $p < .05$. SD, Standard deviation; m, meter; s: second.

$^{\alpha}$ p values evaluating the differences for a given mobility measure (self-propulsion or non-propulsion) across the two time points.

$^{\beta}$ p values evaluating the differences between the two subgroups over time.

$^{\gamma}$ p values evaluating the differences between the two mobility measures (self-propulsion vs. non-propulsion) at a given time point.

[a] Possible reasons for missing data: not measured, loss to follow-up, death, participant no longer able to use a manual wheelchair, inability to perform data collection

[b] Daily mean.

**Table S3. Mean wheelchair objective mobility measures – all and propulsion only – and comparison at a given time point (week)**

| | All | | | ≤ 46 years | | | ≥ 47 years | | | | Males | | | Females | | | |
|---|---|---|---|---|---|---|---|---|---|---|---|---|---|---|---|---|---|
| | (n=29) | | | (n=17) | (n=16) | | (n=12) | (n=13) | | | (n=18) | | | (n=11) | | | |
| | Baseline | Follow-Up | | Baseline | Follow-Up | | Baseline | Follow-Up | | | Baseline | Follow-Up | | Baseline | Follow-Up | | |
| | Mean (SD) | | $^{\alpha}p$ | Mean (SD) | | $^{\alpha}p$ | Mean (SD) | | $^{\alpha}p$ | $^{\beta}p$ | Mean (SD) | | $^{\alpha}p$ | Mean (SD) | | $^{\alpha}p$ | $^{\beta}p$ |
| **Missing** [a]**, n (%)** | 10 (34.5) | 9 (31.0) | | 5 (29.4) | 2 (12.5) | | 5 (41.7) | 7 (53.8) | | | 6 (33.3) | 3 (16.7) | | 4 (36.4) | 6 (54.5) | | |
| **Total distance**, m [b] | | | | | | | | | | | | | | | | | |
| Week – Propulsion | 417.2 (375.5) | 329.7 (239.6) | .71 | 548.8 (402.6) | 393.5 (249.9) | .10 | 191.4 (177.8) | 181.0 (133.6) | .66 | .23 | 491.1 (412.3) | 345.4 (266.8) | .28 | 290.4 (285.7) | 282.6 (140.8) | .70 | .48 |
| Week – Non-propulsion | 386.4 (260.6) | 373.4 (324.3) | .60 | 318.7 (179.2) | 396.6 (379.8) | .69 | 502.6 (346.3) | 319.2 (139.9) | .15 | --- | 398.7 (289.2) | 399.6 (361.0) | .65 | 365.5 (222.8) | 294.5 (181.8) | .80 | --- |
| $^{\gamma}p$ | .89 | .84 | | .12 | .89 | | .07 | .29 | | | .64 | .79 | | .36 | .42 | | |
| Week – All | 803.6 (476.1) | 703.1 (457.1) | .40 | 867.5 (507.7) | 790.0 (520.0) | .51 | 694.1 (430.6) | 500.2 (142.5) | .23 | .67 | 889.8 (511.4) | 745.1 (496.9) | .23 | 655.9 (400.4) | 577.1 (319.2) | .94 | .66 |
| **Number of bouts** [b] | | | | | | | | | | | | | | | | | |
| Week – Propulsion | 45.9 (29.8) | **38.3 (22.3)** | .43 | 47.0 (27.9) | 40.0 (21.3) | .32 | 44.0 (35.1) | 34.1 (26.1) | .93 | .69 | 43.5 (29.8) | 35.8 (20.5) | .56 | 49.9 (31.6) | 45.5 (28.4) | .82 | .86 |
| Week – Non-propulsion | 62.5 (44.0) | **59.9 (41.2)** | .94 | 50.7 (22.3) | 59.3 (46.0) | .47 | 82.8 (64.4) | 61.2 (30.8) | .52 | --- | 65.9 (54.3) | 60.1 (44.8) | .67 | 56.8 (18.5) | 59.1 (32.5) | .48 | --- |
| $^{\gamma}p$ | .14 | **.02** | | .41 | .057 | | .10 | .46 | | | .34 | .12 | | .16 | .17 | | |
| Week – All | 108.4 (64.5) | 98.1 (54.8) | .70 | 97.7 (46.3) | 99.4 (60.4) | .97 | 126.7 (89.1) | 95.3 (43.6) | .38 | .52 | 109.4 (74.5) | 96.0 (55.6) | .51 | 106.7 (48.3) | 104.6 (58.1) | .83 | .73 |
| **Bout length**, m | | | | | | | | | | | | | | | | | |
| Week – Propulsion | 8.4 (7.5) | 7.4 (4.9) | .92 | 11.3 (8.1) | 8.6 (5.3) | .12 | 3.4 (1.8) | 4.7 (2.6) | .23 | .11 | 10.5 (8.7) | 8.2 (5.4) | .41 | 4.7 (2.1) | 5.2 (1.9) | .59 | .45 |
| Week – All | 8.6 (6.6) | 7.8 (5.5) | .69 | 9.7 (7.4) | 8.1 (5.8) | .17 | 6.7 (4.8) | 7.1 (5.1) | .55 | .32 | 9.7 (7.4) | 8.7 (6.0) | .62 | 6.8 (4.8) | 4.9 (1.5) | .61 | .74 |
| **Bout speed**, m/s | | | | | | | | | | | | | | | | | |
| Week – Propulsion | 0.22 (0.14) | 0.19 (0.09) | .94 | 0.27 (0.14) | 0.22 (0.10) | .06 | 0.12 (0.05) | 0.14 (0.05) | .38 | .11 | 0.25 (0.17) | 0.21 (0.10) | .37 | 0.16 (0.05) | 0.15 (0.05) | .81 | .63 |
| Week – All | 0.22 (0.11) | 0.20 (0.11) | .18 | 0.24 (0.12) | 0.21 (0.10) | >.99 | 0.19 (0.09) | 0.20 (0.13) | .93 | .42 | 0.24 (0.12) | 0.22 (0.12) | .17 | 0.20 (0.09) | 0.15 (0.04) | .25 | .67 |
| **Maximum speed**, m/s [b] | | | | | | | | | | | | | | | | | |
| Week – Propulsion | 1.30 (0.59) | 1.12 (0.57) | .16 | **1.58 (0.53)** | **1.29 (0.59)** | **.02** | 0.83 (0.37) | 0.71 (0.23) | .59 | .29 | 1.34 (0.70) | 1.11 (0.60) | .11 | 1.25 (0.40) | 1.15 (0.54) | .94 | .59 |
| Week – All | 1.69 (0.64) | 1.48 (0.61) | .14 | **1.89 (0.70)** | **1.56 (0.67)** | **.04** | 1.35 (0.32) | 1.30 (0.46) | .82 | .50 | 1.70 (0.74) | 1.55 (0.64) | .38 | 1.67 (0.46) | 1.26 (0.51) | .11 | .58 |

All p values obtained using nparLD ANOVAs. Significant at $p < .05$. SD, Standard deviation; m, meter; s: second.

$^{\alpha}$ p values evaluating the differences for a given mobility measure (self-propulsion or non-propulsion) across the two time points.

$^{\beta}$ p values evaluating the differences between the two subgroups over time.

$^{\gamma}$ p values evaluating the differences between the two mobility measures (self-propulsion vs. non-propulsion) at a given time point.

[a] Possible reasons for missing data: not measured, loss to follow-up, death, participant no longer able to use a manual wheelchair, inability to perform data collection

[b] Daily mean.
